# Is aging acceleration mediating the association between hemoglobin glycation index and cardiovascular disease?

**DOI:** 10.1101/2024.07.11.24310308

**Authors:** Na Liu, Yun Li, Mengying Li, Yi Wang, Bo Li, Yongqiang Lian, Jianfang Fu, Xiaomiao Li, Jie Zhou

## Abstract

**Background:** The potential factors beyond HbA1c that increase the risk of cardiovascular disease and age more quickly in people with diabetes are not yet clear. This study sought to determine the prospective associations between discrepancies in observed and predicted HbA1c levels, also known as the hemoglobin glycation index (HGI), and cardiovascular disease risk. Additionally, the interactions of HGI with accelerated aging in relation to cardiovascular disease risk were evaluated.

**Method:** This cross-sectional study included 9167 adults from the National Health and Nutrition Examination Survey 1999–2010. The HGI is used to assess individual blood glucose variability, and phenotypic age acceleration is employed to evaluate accelerated aging. Regression analysis, restricted cubic spline and mediation analysis explore the potential roles of phenotypic age acceleration in the relationship between HGI and CVD mortality.

**Results:** Among the 9167 eligible participants (aged 20 years or older), 4390 (47.9%) were males, and the median (IQR) age was 48.0 (15.0) years; 4403 (48.0%) had prediabetes and diabetes, and 985 (10.7%) had cardiovascular disease. Restricted cubic splines showed that the association between HGI and CVD risk was nonlinear (p < 0.001). The greater the negative value of the HGI was, the greater the risk of CVD, and the association was independent of age, sex and HbA1c. Mediation analyses confirmed that phenotypic age acceleration acted as a mediator in the association between HGI and CVD risk (mediated effect: OR, 68.7%, 95% CI: 36.4%-153%, *P*=0.002).

**Conclusion and Relevance:** The HGI serves as a robust biomarker for assessing the acceleration of aging, regardless of HbA1c levels, and is associated with increased susceptibility to cardiovascular disease, particularly among individuals characterized by negative HGI.

## Introduction

Type 2 diabetes (T2D) and prediabetes are prevalent in individuals with cardiovascular disease (CVD) and are linked to adverse cardiovascular outcomes^1^. Glycated hemoglobin A1c (HbA1c), a marker of protein glycosylation in erythrocytes, can partially indicate cardiovascular disease risk in people with diabetes^2^. However, it is crucial to acknowledge that HbA1c inadequately represents glycemic control in approximately 40% of individuals with diabetes^3^. Hence, identifying additional factors influencing CVD risk beyond HbA1c may help mitigate the impact of this condition on adults, particularly in individuals with diabetes.

Elevated glucose concentrations exacerbate the aging process and contribute to cardiac damage. The hemoglobin glycation index (HGI), a measure of the mismatch between observed and predicted HbA1c, reflects interindividual glucose variability^3^. Strong evidence indicates that the HGI can serve as an indicator of vascular health status^4^ and is associated with the risk of various diabetic vascular complications. The exact mechanisms mediating this association remain unknown, raising the question of whether aging acceleration could play a prominent role. Aging markers in clinical practice, such as leukocyte telomere length (LTL)^5^ and phenotypic age acceleration (PAA)^6^, have been correlated with CVD. We hypothesized that aging acceleration could be a mediator between HGI and CVD risk. Therefore, the detection of a significantly lower HbA1c than blood sugar-matched HbA1c in patients may increase the risk of age-related diseases, such as CVD. Accordingly, the primary objective of this study was to investigate the impact of HGI on CVD using NHANES surveys of US populations while also ascertaining the influence of aging on this process.

## Methods

### Study population

The NHANES is an ongoing national cross-sectional survey, and data are available on the website of the American Centers for Disease Control and Prevention (http://www.cdc.gov/nchs/nhanes.htm). The study protocol was approved by the National Center for Health Statistics’ ethics review committee. All participants provided written consent at the time of recruitment. The NHANES 1999-2010 data were chosen because C-reactive protein (CRP), which was used to calculate phenotypic age, was collected from these years.

A total of 59,731 adults (aged 20 to 85 years) were surveyed. Next, we excluded individuals with missing laboratory data regarding age, albumin, creatinine, fasting plasma glucose (FPG), CRP, alkaline phosphatase, and complete blood count with 5-part differential (n=46,958), which were used to calculate phenotypic age; missing data on HbA1c (n=26), which were used to calculate HGI; and missing questionnaire data about CVD (n=3,580). All participants were free of severe anemia. Overall, a total of 9167 adults were surveyed, including 985 CVD patients.

### Disease ascertainment

The diagnosis of CVD was assessed by the NHANES medical conditions questionnaire (MCQ): “Has a doctor or other health expert ever informed you that you have congestive heart failure/coronary heart disease/angina pectoris/ myocardial infarction /stroke (MCQ160B-F)?” A person was regarded as having CVD if he or she replied “yes” to any of the above questions. Participants who provided “Do not know” or “Refused” responses were considered to have missing data.

According to the ADA’s diabetes diagnostic criteria^7^, diagnosed diabetes and prediabetes were defined by self-reported diagnosis and met 1 of the laboratory criteria (FPG level of ≥ 5.6 mmol/l or HbA1c level ≥ 5.7%).

### Key variables

#### (1) Biological aging markers

Phenotypic age was calculated by using 9 aging-related variables^8^, including chronological age, albumin, creatinine, glucose, CRP, lymphocyte percentage, mean cell volume, red blood cell distribution width, alkaline phosphatase, and white blood cell count.

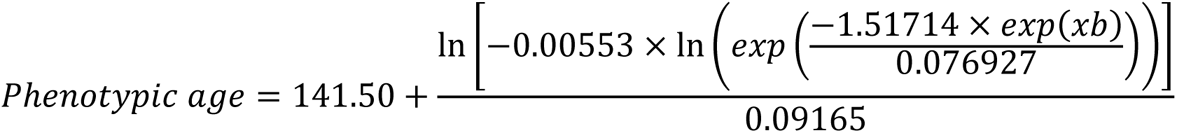

where

xb = − 19.907 − 0.0336 × Albumin + 0.0095 × Creatinine + 0.1953 × Glucose + 0.0954 × LnC RP-0.0120 × Lymphocyte Percent + 0.0268 × Mean Cell Volume + 0.3306 × Red Cell Distribution Width + 0.00188 × Alkaline Phosphatase + 0.0554 × White Blood Cell Count + 0.0804 × Chronological Age.

We then estimated PAA, calculated as the residuals resulting from a linear model regressing phenotypic age on chronological age. A negative PAA value (PAA<0) indicated a slow aging group, indicating that the phenotypic age was younger than the chronological age. Conversely, a positive PAA value (PAA≥0) indicates accelerated aging.

#### (2) Telomere length

We combined the NHANES surveys conducted in 1999-2000 and 2001-2002 because these surveys measure LTL. The full details are available at http://cdc.gov/nchs/nhanes. The length of telomeres is represented by the mean (standard deviation) of the telomere length relative to the standard reference DNA ratio.

#### (3) HGI

The mismatch between glycemia and HbA1c was measured by the HGI. First, linear regression models of observed HbA1c vs. observed FPG were used. The association of HbA1c levels with FPG levels varied according to y-intercept. predicted HbA1c= (0.45*FPG) + 2.99. HGI = observed-predicted HbA1c. Next, the HGI was categorized into tertiles. Participants in the highest tertile of HGI exhibited an observed HbA1c level that exceeded the predicted value. Those in the moderate tertile had an observed HbA1c close to the predicted level, resulting in a near-zero HGI. Participants in the lowest tertile of HGI demonstrated an observed HbA1c lower than predicted, leading to a negative HGI value.

### Statistical analysis

All the statistical analyses were performed using R version 4.1.2. Missing data were imputed using the multiple imputation chained equation procedure. Continuous variables are reported as either the mean (±SD, standard deviation) or median (interquartile range [IQR]), while categorical variables are presented as frequencies and percentages. Continuous variables were analyzed by HGI tertiles using ANOVA, while categorical variables were assessed using chi-square tests.

Multivariate linear regression models were developed to investigate the association between HGI and accelerated aging, and β coefficients and their corresponding standard errors were calculated. Multivariate logistic regression models were then constructed to explore potential associations between HGI and CVD incidence, along with odds ratios (ORs) and 95% confidence intervals (CIs). PAA was used as a possible mediator of the association between HGI and CVD incidence. We separately used linear regression and logistic regression to examine the relationships between HGI and PAA and between HGI and CVD mortality, with a 4-knot (5th, 35th, 65th, 95th percentiles of HGI) (0.05, 0.35, 0.65, 0.95) restricted cubic spline and smooth curve fitting. To test the robustness and potential variations in different subgroups, we repeated the analyses stratified by sex (men and women), glucose state (NGM, prediabetes and diabetes). P < 0.05 was considered to indicate statistical significance.

## Results

### 1. General characteristics of participants according to HGI tertiles

The general characteristics of participants from the US NHANES by tertiles of HGI are shown in Table 1. This study included 9167 participants with a median (IQR) age of 48.00 (15) years and 48% males. Compared with those in tertile 1, participants in the other tertiles were more likely to be older, female, and have a higher body mass index (BMI), HbA1c, TC, and LDL. Specifically, participants in tertile 2 (moderate HGI) tended to have lower FPG, homeostasis model assessment-insulin resistance (HOMA-IR) levels and prevalence rates of prediabetes and diabetes.

**Table 1.**
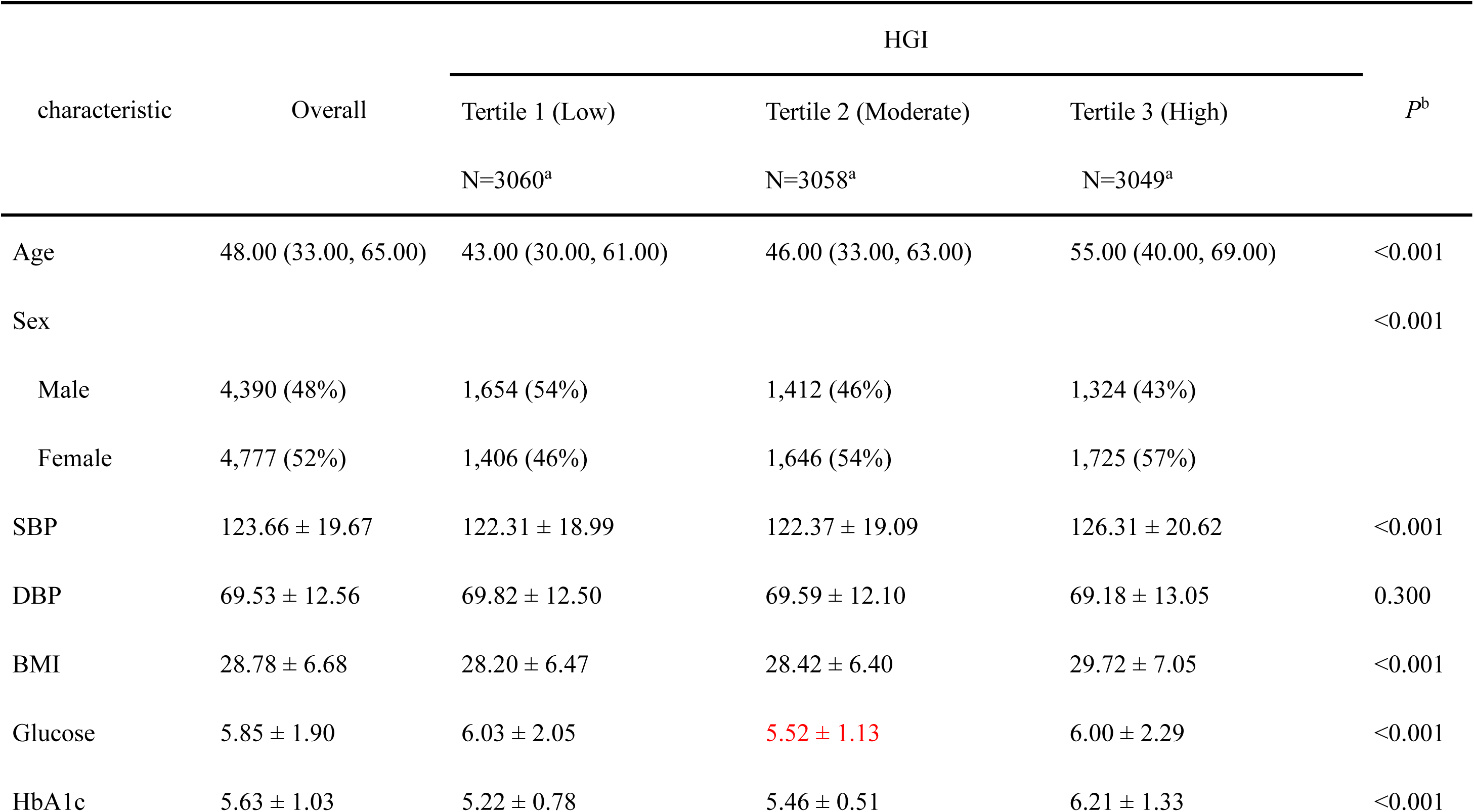
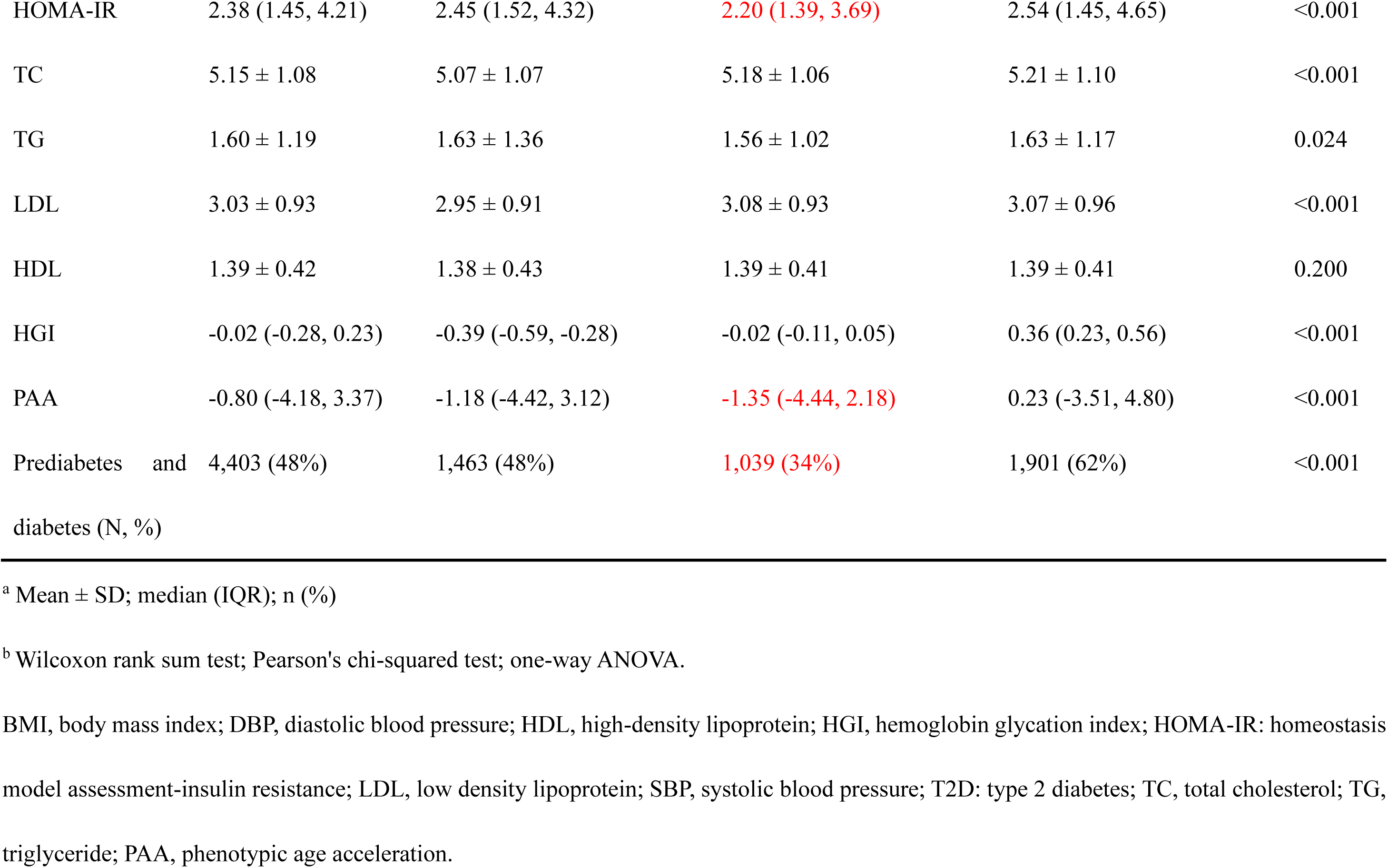
Baseline characteristics by HGI among participants in the NHANES.

| characteristic | Overall | HGI |  |  | *P*<sup>b</sup> |
| --- | --- | --- | --- | --- | --- |
|  |  | Tertile 1 (Low)<br>N=3060<sup>a</sup> | Tertile 2 (Moderate)<br>N=3058<sup>a</sup> | Tertile 3 (High)<br>N=3049<sup>a</sup> |  |
| Age | 48.00 (33.00, 65.00) | 43.00 (30.00, 61.00) | 46.00 (33.00, 63.00) | 55.00 (40.00, 69.00) | <0.001 |
| Sex |  |  |  |  | <0.001 |
| Male | 4,390 (48%) | 1,654 (54%) | 1,412 (46%) | 1,324 (43%) |  |
| Female | 4,777 (52%) | 1,406 (46%) | 1,646 (54%) | 1,725 (57%) |  |
| SBP | 123.66 ± 19.67 | 122.31 ± 18.99 | 122.37 ± 19.09 | 126.31 ± 20.62 | <0.001 |
| DBP | 69.53 ± 12.56 | 69.82 ± 12.50 | 69.59 ± 12.10 | 69.18 ± 13.05 | 0.300 |
| BMI | 28.78 ± 6.68 | 28.20 ± 6.47 | 28.42 ± 6.40 | 29.72 ± 7.05 | <0.001 |
| Glucose | 5.85 ± 1.90 | 6.03 ± 2.05 | 5.52 ± 1.13 | 6.00 ± 2.29 | <0.001 |
| HbA1c | 5.63 ± 1.03 | 5.22 ± 0.78 | 5.46 ± 0.51 | 6.21 ± 1.33 | <0.001 |
| HOMA-IR | 2.38 (1.45, 4.21) | 2.45 (1.52, 4.32) | 2.20 (1.39, 3.69) | 2.54 (1.45, 4.65) | <0.001 |
| TC | 5.15 ± 1.08 | 5.07 ± 1.07 | 5.18 ± 1.06 | 5.21 ± 1.10 | <0.001 |
| TG | 1.60 ± 1.19 | 1.63 ± 1.36 | 1.56 ± 1.02 | 1.63 ± 1.17 | 0.024 |
| LDL | 3.03 ± 0.93 | 2.95 ± 0.91 | 3.08 ± 0.93 | 3.07 ± 0.96 | <0.001 |
| HDL | 1.39 ± 0.42 | 1.38 ± 0.43 | 1.39 ± 0.41 | 1.39 ± 0.41 | 0.200 |
| HGI | -0.02 (-0.28, 0.23) | -0.39 (-0.59, -0.28) | -0.02 (-0.11, 0.05) | 0.36 (0.23, 0.56) | <0.001 |
| PAA | -0.80 (-4.18, 3.37) | -1.18 (-4.42, 3.12) | -1.35 (-4.44, 2.18) | 0.23 (-3.51, 4.80) | <0.001 |
| Prediabetes and diabetes (N, %) | 4,403 (48%) | 1,463 (48%) | 1,039 (34%) | 1,901 (62%) | <0.001 |

<sup>a</sup> Mean ± SD; median (IQR); n (%)
<sup>b</sup> Wilcoxon rank sum test; Pearson's chi-squared test; one-way ANOVA.
BMI, body mass index; DBP, diastolic blood pressure; HDL, high-density lipoprotein; HGI, hemoglobin glycation index; HOMA-IR: homeostasis model assessment-insulin resistance; LDL, low density lipoprotein; SBP, systolic blood pressure; T2D: type 2 diabetes; TC, total cholesterol; TG, triglyceride; PAA, phenotypic age acceleration.

### 2. Associations between HGI and total CVD incidence

Among the 9167 participants, we identified 985 individuals with CVD. The subgroup analysis of logistic regression showed that HGI was not an independent risk factor for CVD in the moderate-HGI group. With adjustment for sex and age, the larger the absolute value of the HGI was, the greater the risk of CVD. Interestingly, after further controlling for HbA1c, positive HGI was no longer a risk factor for CVD, while negative HGI remained a risk factor for CVD (Table 2). According to the restricted cubic spline regression models, the associations of HGI and combined HbA1c with CVD incidence are shown in Fig. 1 and Figure S1. There was a U-shaped relationship between HGI and CVD risk; however, there was an approximate L-shaped relationship between HGI and CVD risk after further controlling for HbA1c.

**Figure 1.**
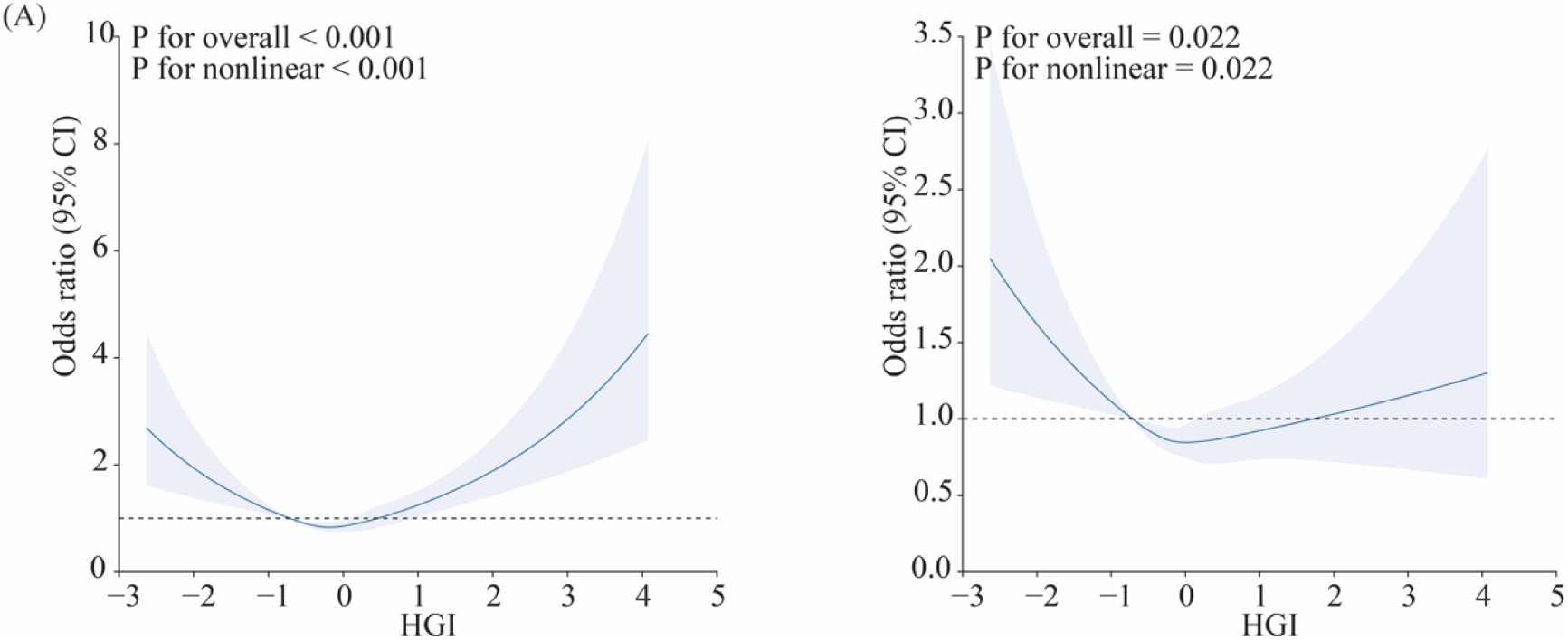
Association Between HGI and CVD Using a Restricted Cubic Spline Regression Model. Graphs show ORs for CVD according to HGI adjusted for age and sex (A) and age, sex, and HbA1c (B). Data were fitted by a logistic regression model, and the model was conducted with 4 knots at the 5th, 35th, 65th, 95th percentiles of HGI (reference is the 5th percentile). Solid lines indicate ORs, and shadow shape indicate 95% CIs. OR, odds ratio; CI, confidence interval.

**Table 2.** Logistic regression analysis for the association between HGI and cardiovascular disease.

|  | Crude |  | Model 1 |  | Model 2 |  |
| --- | --- | --- | --- | --- | --- | --- |
|  | OR (95% CI) | *P* | OR (95% CI) | *P* | OR (95% CI) | *P* |
| Low HGI, N=3060 | 0.454 (0.351-0.588) | <0.001 | 0.574 (0.441-0.746) | <0.001 | 0.623 (0.471-0.824) | 0.001 |
| Moderate HGI, N=3058 | 1.493 (0.417-5.348) | 0.538 | 1.176 (0.296-4.679) | 0.818 | 0.967 (0.241-3.872) | 0.962 |
| High HGI, N=3049 | 1.593 (1.373-1.8848) | <0.001 | 1.579 (1.340-1.861) | <0.001 | 1.177 (0.941-1.472) | 0.153 |

CI, confidence interval; HGI, hemoglobin glycation index; OR, odds ratio.
Model 1, adjusted for sex and age; Model 2, adjusted for sex, age, and HbA1c.

### 3. Relationship between HGI and accelerated aging

Apart from glycemic control, the duration of erythrocyte survival significantly impacts HbA1c levels. There was a significant correlation between HbA1c and HGI in the accelerated aging group (r = 0.8, *P*<0.001), which was significantly greater than that in the decelerated aging group (Figure S2A). We conducted further analysis to investigate the association between HGI and aging. The general characteristics of the participants with LTL (NHANES 1999-2012) are shown in Table S1. First, there is an inverted U-shaped relationship between HGI and LTL. The absolute magnitude of HGI was negatively associated with telomere length (Figure S2B).

Additionally, we employed a restricted cubic spline to flexibly model and visualize the associations between HGI, HbA1c and PAA (Fig. 2). A V-shaped correlation was observed between HGI and PAA (p nonlinear 0.002). There was a stronger correlation between HGI and PAA in people with diabetes or prediabetes than in those with NGM (Figure S2C-D). After adjusting for HbA1c, a robust negative linear correlation between HGI and PAA was observed, confirming that positive HGI is influenced mainly by HbA1c. Conversely, negative HGI remains significantly associated with accelerated aging.

**Figure 2.**
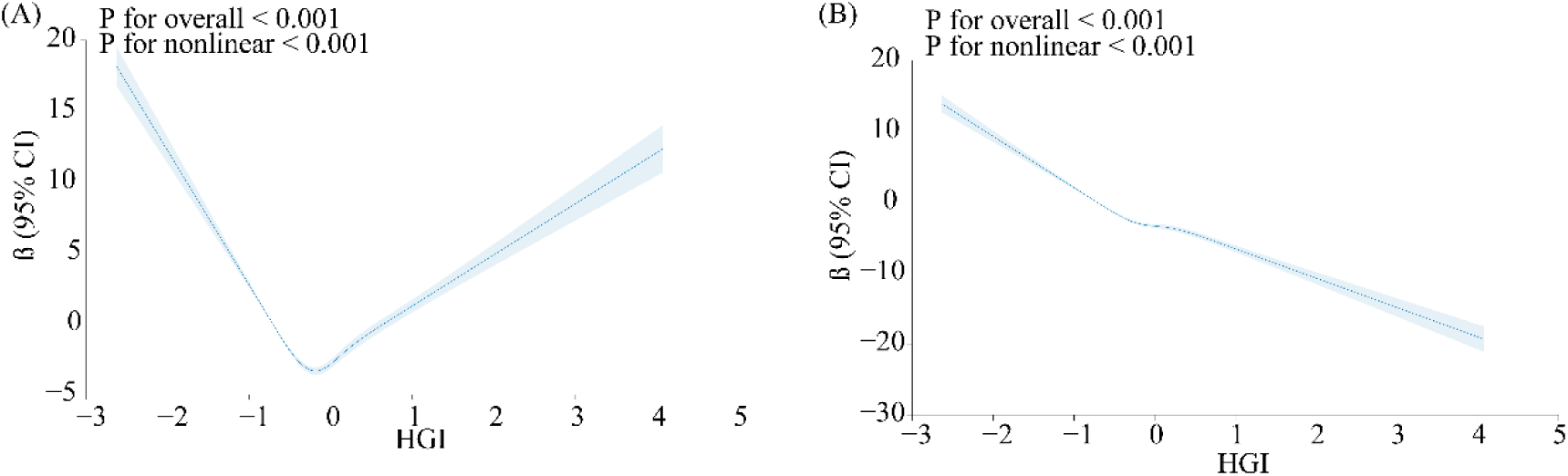
Association Between HGI and PAA Using a Restricted Cubic Spline Regression Model. Graphs show β for PAA according to HGI (A) and adjusted for HbA1c (B). Data were fitted by a linear regression model, and the model was conducted with 4 knots at the 5th, 35th, 65th, 95th percentiles of HGI (reference is the 5th percentile). Solid lines indicate β, and shadow shape indicate 95% CIs. CI, confidence interval.

### 4. The modifying effect of aging acceleration on the association between HGI and CVD risk

The length of telomeres is inversely associated with the risk of CVD, whereas elevated levels of PAA are positively associated with the risk of CVD (Figure S4A-B). The association between negative HGI and CVD risk disappeared after further controlling for PAA compared to solely controlling for sex and age (Fig. 3A). Mediation analysis was used to investigate the mediating effect of PAA between negative HGI and CVD. Specifically, HGI was negatively correlated with PAA (β ± SE = −8.456 ± 0.3369, p < 0.001). Ultimately, the association between HGI and CVD incidence was mediated by PAA in 68.7% (95% CI 36.4%-153%) of the observational studies (Fig. 3B).

**Figure 3.**
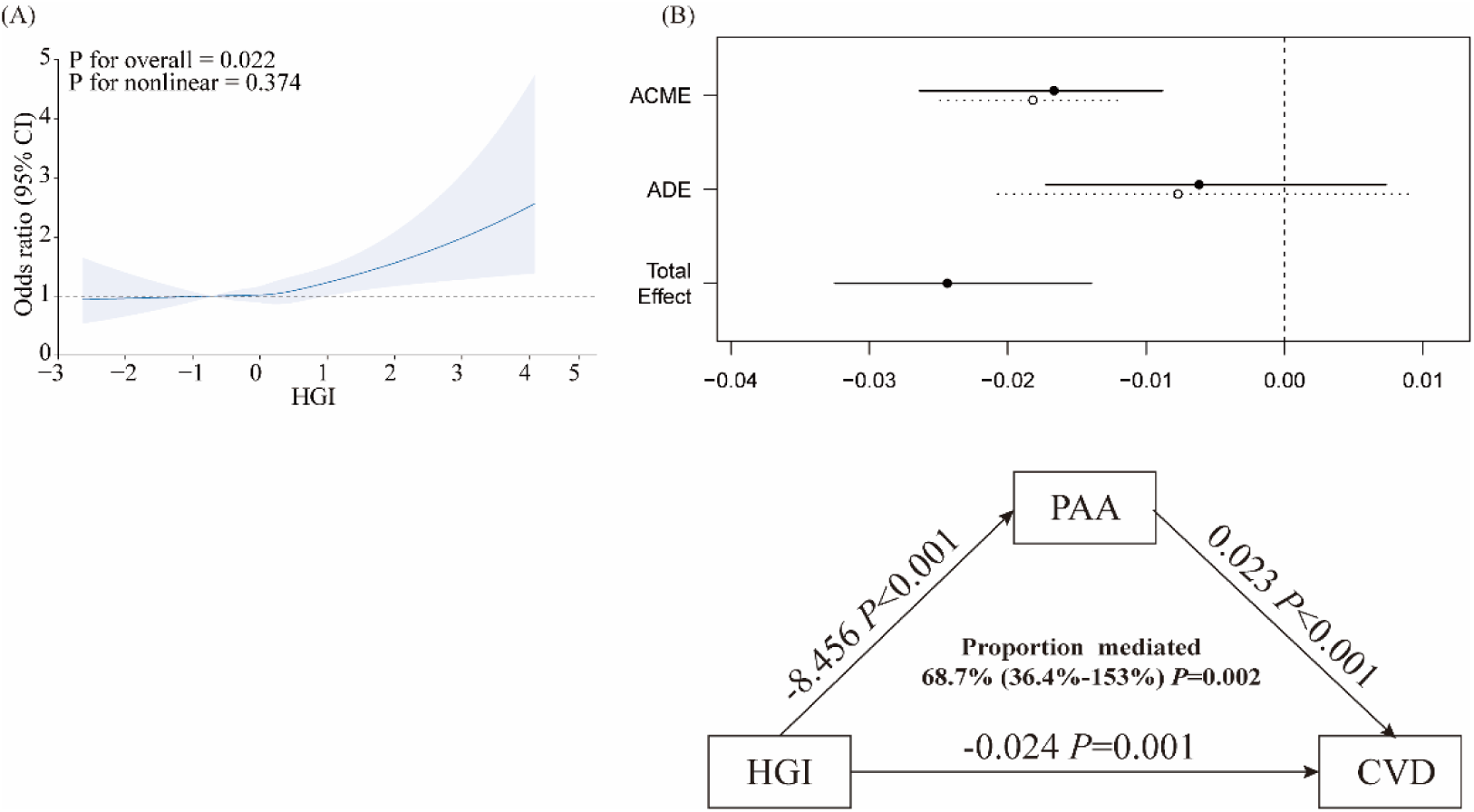
The relationship between negative HGI and CVD risk is mediated by accelerated aging. Graphs show ORs for CVD according to HGI (A) and mediating effect of PAA on the relationship between HGI and the CVD risk in HGI tertile 1 (B). Adjusted for age, sex, and PAA. ACME, Average Causal Mediation Effect; ADE, Average Direct Effect.

## Discussion

This study generated two key findings. First, we found that a negative HGI, rather than a positive HGI, was significantly associated with a greater risk of CVD independent of HbA1c, particularly in people with diabetes. Additionally, we observed that a negative HGI is a potent and likely neglected indicator of accelerated aging, and identifying individuals with negative HGI is crucial for the prevention and early intervention of aging-related diseases such as CVD.

Diabetes is widely accepted as a risk factor for CVD. HbA1c, as a *sine qua non* of diabetes diagnosis, cannot capture glycemic variations well concentrations. In clinical practice, there is often a discrepancy between plasma glucose and HbA1c levels, with the mismatch becoming more prevalent in individuals with higher blood glucose concentrations. The HGI reflects the difference between the predicted and observed HbA1c. People with positive HGI results were mainly elderly people with diabetes and prediabetes, and people with negative HGI results were mainly people with NGM. Individuals with positive HGI are prone to receiving excessive and inappropriate treatment for hypoglycemia, while those with negative HGI experience suboptimal glycemic control and an increased risk of diabetes-related complications^3^. Many doubts have been aired about the consistent discrepancy between glycated hemoglobin and actual glycemia. The current consensus is lacking regarding whether HGI, as an indicator of individual plasma glucose variability, can effectively reflect the variations in CVD risk among individuals with diabetes. An increasing body of evidence indicates that a positive HGI is associated with increased protein glycation, can serve as an indicator of vascular health status in individuals with impaired glucose metabolism^4^ and is associated with the risk of various diabetic vascular complications, including CVD^4^ and diabetic nephropathy^9^. However, certain studies have proposed that the association between HGI and macrovascular complications diminishes after adjusting for HbA1c^10^. To elucidate the relationship between HGI and the occurrence of CVD, we carried out a cross-sectional analysis of 9167 participants from the NHANES cohort. The present study revealed that individuals with abnormal sugar metabolism exhibit a greater degree of variability in HGI than do those with NGM. A positive HGI heightens susceptibility to CVD, which is well reported^11^. There is limited literature available on the association between negative HGI and CVD risk. The present study revealed that negative HGI levels were associated with a greater risk of CVD in individuals with normoglycemia and abnormal glucose metabolism, indicating a U-shaped relationship consistent with previous studies^12^. This U-shaped relationship between HGI and CVD risk also holds true in individuals with NGM but exhibits a stronger association with impaired glucose regulation.

Although the precise biological mechanism underlying the correlation between HGI and CVD risk remains unclear, it may be closely related to HbA1c, and previous studies have not assessed this correlation. Our results suggest an approximate inverse correlation between HGI and the incidence of CVD after adjusting for HbA1c. The HbA1c level is influenced by the lifespan of red blood cells ^13^ and the glucose gradient across their cell membrane^14^. The longer the average lifespan of red blood cells is, the greater the measured level of HbA1c; conversely, the shorter the average lifespan of red blood cells is, the lower the measured level of HbA1c^13^. This inaccurately reflects actual blood glucose concentrations over the previous 2-3 months. However, certain studies have proposed that there is considerable variation in red blood cell survival rates within normal populations, with both individuals with DM and those without DM having similar average lifespans and ranges for erythrocytes^13^. Therefore, can the association between HGI and CVD risk be elucidated by examining cellular senescence?

Diabetes, along with other metabolic disorders, is commonly regarded as a condition that accelerates aging. The risk of myocardial damage in diabetic patients is significantly increased, which can be attributed not only to elevated blood sugar levels but also to aging^15^. The rate of aging varies among individuals, resulting in differences in susceptibility to CVD as individuals age^16^. Therefore, distinguishing the aging process among individuals of the same chronological age will enhance early prevention by enabling earlier identification of individuals at high risk for CVD. Biological aging is known to be assessed in a variety of ways, with the most prominent ones being epigenetic clocks (represented as DNA methylation age) and the lLTL^17^. Recently, aging measures based on clinically observable data, such as PAA, have been shown to strongly predict susceptibility to various metabolic diseases^18^. Our study evaluated the relationship between CVD and biological aging and demonstrated a significant positive association between accelerated aging and CVD risk, in line with previous findings ^6,19^. .

To further elucidate whether HGI can accurately reflect interindividual differences in aging, we analyzed the relationship between HGI and PAA. The results indicated a U-shaped relationship between HGI and PAA, with lower PAA observed when HGI was close to zero. This suggests that a closer alignment between the predicted and measured HbA1c levels corresponds to a slower aging process, whereas a larger disparity indicates accelerated aging. After controlling for HbA1c, HGI showed a nearly linear negative correlation with PAA, suggesting that the relationship between HGI and aging is influenced by HbA1c. The relationship between PAA and CVD risk in relation to HGI was further assessed in this study. Notably, our findings demonstrate for the first time that the association between negative HGI levels and increased CVD risk is no longer observed after adjusting for PAA. Mediation analysis suggested that the relationship between negative HGI and CVD is fully mediated by PAA. Previous studies have suggested that the HGI reflects interindividual variability in susceptibility to hemoglobin glycation. Based on our findings, we propose that the discrepancy between the measured HbA1c and true glucose concentrations may be attributed to accelerated physiological aging. The variation in HGI is predominantly associated with the aging process and may serve as an indicator of the aging rate to a certain extent. Consequently, the relationship between HGI and cardiovascular disease risk can be elucidated by accelerated physiological aging.

Telomeres, which are indicators of bodily aging, are closely associated with the occurrence of age-related diseases such as metabolic syndrome, T2D, and CVD^20^. The LTL serves as a more accurate reflection of aging and can predict the risk of age-related diseases such as CVD^21,22^. To further elucidate the relationship between HGI and aging, we explored the correlation between HGI and telomere length. Our results revealed a negative correlation between LTL and CVD, providing evidence that CVD is an age-related disease. The absolute magnitude of HGI, indicating the discrepancy between predicted and actual HbA1c levels, is positively associated with telomere shortening. Previous studies have shown that the moderate-HGI group exhibited the highest mtDNAcn, while the high-HGI group showed a decrease without statistically significant differences^23^, which indirectly suggested that both excessively positive and negative HGIs are associated with accelerated aging. Our results provide additional evidence supporting a positive correlation between the absolute magnitude of HGI and telomere shortening. Elevated glycemia can lead to reduced activity of erythrocyte Na^+^-K^+^- ATPase, resulting in an increased glucose gradient on the red blood cell membrane^24^ and increased HGI. Moreover, aging may contribute to a decrease in erythrocyte ATP content^25^. The interaction between diabetes and aging is associated with an increased risk of complications such as CVD. Overall, HGI is linked to accelerated aging and shortened telomeres.

The HGI is a clinically validated measurement index that is easily available in routine clinical practice. Therefore, the HGI can serve as a convenient and accessible indicator of CVD risk to assist clinicians in monitoring the risk of CVD in individuals, especially those with prediabetes or diabetes. However, the lack of OGTT data might lead to potential underdiagnosis. Second, the assessment of CVD risk was based on self-report questionnaires, which are susceptible to recall bias. Due to the cross-sectional design of this study, it is challenging to establish a causal relationship between HGI and CVD. Further research is required to comprehensively understand the mechanisms underlying this correlation and whether targeting cellular aging pathways could reduce the risk of CVD in individuals with elevated HGI levels.

## Conclusion

The study presented evidence that individuals with a negative HGI may be at an elevated risk for cardiovascular disease. HGI, being a clinically accessible and valuable indicator, can be utilized to assess the acceleration of aging and is closely associated with an increased susceptibility to age-related health issues, including cardiovascular disease. These findings have the potential to aid in identifying individuals who could benefit from targeted interventions, particularly in T2D, aimed at reducing their risk of developing cardiovascular disease as they age.

## Data Availability

Data is available on the website of the America Centers for Disease Control and Prevention (CDC)

http://www.cdc.gov/nchs/nhanes.htm

## Acknowledgements

We sincerely thank all individuals who contributed to the study of NHANES.

## Sources of Funding

National Natural Science Foundation of China (NSFC): 82070839

## Disclosures

None.

## Notes

### Competing Interest Statement

The authors have declared no competing interest.

### Clinical Trial

The analyses utilized existing data from the NHANES study, and we were not involved in participant recruitment.

### Author Declarations

Ethics Review Board of the National Center for Health Statistics

## References

1. Marx N, Federici M, Schütt K, Müller-Wieland D, Ajjan RA, Antunes MJ, Christodorescu RM, Crawford C, Di Angelantonio E, Eliasson B, et al. 2023 ESC Guidelines for the management of cardiovascular disease in patients with diabetes. Eur. Heart J. 2023;44:4043–4140.

2. Cahn A, Wiviott SD, Mosenzon O, Goodrich EL, Murphy SA, Yanuv I, Rozenberg A, Bhatt DL, Leiter LA, McGuire DK, et al. Association of Baseline HbA1c With Cardiovascular and Renal Outcomes: Analyses From DECLARE-TIMI 58. Diabetes Care. 2022;45:938–946.

3. Nayak AU, Singh BM, Dunmore SJ. Potential Clinical Error Arising From Use of HbA1c in Diabetes: Effects of the Glycation Gap. Endocr. Rev. 2019;40:988–999.

4. Ahn CH, Min SH, Lee D-H, Oh TJ, Kim KM, Moon JH, Choi SH, Park KS, Jang HC, Ha J, et al. Hemoglobin Glycation Index Is Associated With Cardiovascular Diseases in People With Impaired Glucose Metabolism. J. Clin. Endocrinol. Metab. 2017;102:2905– 2913.

5. Schuermans A, Nakao T, Uddin MM, Hornsby W, Ganesh S, Shadyab AH, Liu S, Haring B, Shufelt CL, Taub MA, et al. Age at Menopause, Leukocyte Telomere Length, and Coronary Artery Disease in Postmenopausal Women. Circ. Res. 2023;133:376–386.

6. Chen L, Yin X, Zhao Y, Chen H, Tan T, Yao P, Tang Y. Biological ageing and the risks of all-cause and cause-specific mortality among people with diabetes: a prospective cohort study. J. Epidemiol. Community Health. 2022;76:771–778.

7. American Diabetes Association. 2. Classification and Diagnosis of Diabetes: Standards of Medical Care in Diabetes-2020. Diabetes Care. 2020;43:S14–S31.

8. Levine ME, Lu AT, Quach A, Chen BH, Assimes TL, Bandinelli S, Hou L, Baccarelli AA, Stewart JD, Li Y, et al. An epigenetic biomarker of aging for lifespan and healthspan. Aging. 2018;10:573–591.

9. Xin S, Zhao X, Ding J, Zhang X. Association between hemoglobin glycation index and diabetic kidney disease in type 2 diabetes mellitus in China: A cross-sectional inpatient study. Front. Endocrinol. 2023;14:1108061.

10. Rhee E-J, Cho J-H, Kwon H, Park SE, Park C-Y, Oh K-W, Park S-W, Lee W-Y. Association Between Coronary Artery Calcification and the Hemoglobin Glycation Index: The Kangbuk Samsung Health Study. J. Clin. Endocrinol. Metab. 2017;102:4634–4641.

11. Brownlee M. Biochemistry and molecular cell biology of diabetic complications. Nature. 2001;414:813–820.

12. J Y, Q S, G X, M Y, G S. Sex-specific associations between haemoglobin glycation index and the risk of cardiovascular and all-cause mortality in individuals with pre-diabetes and diabetes: A large prospective cohort study. Diabetes Obes. Metab. [Internet]. 2024 [cited 2024 Jun 19];26. Available from: https://pubmed.ncbi.nlm.nih.gov/38454654/

13. Cohen RM, Franco RS, Khera PK, Smith EP, Lindsell CJ, Ciraolo PJ, Palascak MB, Joiner CH. Red cell life span heterogeneity in hematologically normal people is sufficient to alter HbA1c. Blood. 2008;112:4284–4291.

14. Khera PK, Joiner CH, Carruthers A, Lindsell CJ, Smith EP, Franco RS, Holmes YR, Cohen RM. Evidence for interindividual heterogeneity in the glucose gradient across the human red blood cell membrane and its relationship to hemoglobin glycation. Diabetes. 2008;57:2445–2452.

15. Chia CW, Egan JM, Ferrucci L. Age-related Changes in Glucose Metabolism, Hyperglycemia, and Cardiovascular Risk. Circ. Res. 2018;123:886.

16. L L, L B, F M, Tf L, P L, Gg C. Inflammation, Aging, and Cardiovascular Disease: JACC Review Topic of the Week. J. Am. Coll. Cardiol. [Internet]. 2022 [cited 2024 Jun 19];79. Available from: https://pubmed.ncbi.nlm.nih.gov/35210039/

17. R D, Q F, Y S, Q L. Epigenetic clock: A promising biomarker and practical tool in aging. Ageing Res. Rev. [Internet]. 2022 [cited 2024 Jun 19];81. Available from: https://pubmed.ncbi.nlm.nih.gov/36206857/

18. Liu Z, Kuo P-L, Horvath S, Crimmins E, Ferrucci L, Levine M. A new aging measure captures morbidity and mortality risk across diverse subpopulations from NHANES IV: A cohort study. PLoS Med. 2018;15:e1002718.

19. M C-H, B B, R V, M K, V Z, R C, J E, D M, D P, M W, et al. Education, biological ageing, all-cause and cause-specific mortality and morbidity: UK biobank cohort study. EClinicalMedicine [Internet]. 2020 [cited 2024 Jun 19];29–30. Available from: https://pubmed.ncbi.nlm.nih.gov/33437953/

20. Cheng F, Carroll L, Joglekar MV, Januszewski AS, Wong KK, Hardikar AA, Jenkins AJ, Ma RCW. Diabetes, metabolic disease, and telomere length. Lancet Diabetes Endocrinol. 2021;9:117–126.

21. Cheng F, Luk AO, Tam CHT, Fan B, Wu H, Yang A, Lau ESH, Ng ACW, Lim CKP, Lee HM, et al. Shortened Relative Leukocyte Telomere Length Is Associated With Prevalent and Incident Cardiovascular Complications in Type 2 Diabetes: Analysis From the Hong Kong Diabetes Register. Diabetes Care. 2020;43:2257–2265.

22. Peng H, Mete M, Desale S, Fretts AM, Cole SA, Best LG, Lin J, Blackburn E, Lee ET, Howard BV, et al. Leukocyte telomere length and ideal cardiovascular health in American Indians: the Strong Heart Family Study. Eur. J. Epidemiol. 2017;32:67–75.

23. L L, J Y, Y L, S H, Y Z, M Q, F P, L X, W L, H Z, et al. High Hemoglobin Glycation Index Is Associated With Telomere Attrition Independent of HbA1c, Mediated by TNFα. J. Clin. Endocrinol. Metab. [Internet]. 2022 [cited 2024 Jun 19];107. Available from: https://pubmed.ncbi.nlm.nih.gov/34562085/

24. Fb D, Pj D, G N, Sd B, M M, S G, Mj F. The effect of in vivo glucose administration on human erythrocyte Ca2+-ATPase activity and on enzyme responsiveness in vitro to thyroid hormone and calmodulin. Diabetes [Internet]. 1985 [cited 2024 Jun 19];34. Available from: https://pubmed.ncbi.nlm.nih.gov/2989051/

25. Rabini RA, Petruzzi E, Staffolani R, Tesei M, Fumelli P, Pazzagli M, Mazzanti L. Diabetes mellitus and subjects’ ageing: a study on the ATP content and ATP-related enzyme activities in human erythrocytes. Eur. J. Clin. Invest. 1997;27:327–332.

